# IMPACT OF MEDICAID EXPANSION ON INSURANCE COVERAGE AND COST BARRIERS TO CARE: A DIFFERENCE-IN-DIFFERENCES ANALYSIS USING BRFSS DATA

**DOI:** 10.1101/2025.08.01.25332619

**Authors:** Newton Nyirenda, Bwalya Chisanga, Hannah Muturi, Linda Siachalinga

**Author notes:** Corresponding Author: Newton Nyirenda.

## Abstract

**Background:** The Affordable Care Act (ACA) expanded Medicaid eligibility to improve healthcare access. This study evaluates whether Medicaid expansion led to increased insurance coverage and reduced cost-related barriers to healthcare.

**Methods:** We used Behavioral Risk Factor Surveillance System (BRFSS) data from 2012, 2013, 2015, 2016, and 2018. A Difference-in-Differences (DiD) approach was applied, comparing adults in expansion versus non-expansion states before and after 2014. Logistic regression was employed to calculate odds ratios (OR) along with their corresponding 95% confidence intervals (CI).

**Results:** Medicaid expansion was associated with 25% greater odds of insurance coverage (OR = 1.25; 95% CI: 1.22, 1.28) and 10% lower odds of cost-related barriers to care (OR = 0.90; 95% CI: 0.89 to 0.91, indicating a statistically significant association (p < 0.001).

**Conclusion:** Medicaid expansion under the ACA significantly improved healthcare access by increasing insurance coverage and reducing cost-related barriers.

## INTRODUCTION

Access to affordable healthcare remains a critical challenge in the United States (US). Prior to the implementation of the Affordable Care Act (ACA) in 2010, millions of Americans, particularly those with low incomes, lacked health insurance coverage (1–3). The ACA introduced several reforms aimed at increasing healthcare access, reducing costs, and improving health outcomes. A central component of the ACA involved broadening Medicaid eligibility to include low-income adults earning up to 138% of the federal poverty threshold (1). However, following the 2012 U.S. Supreme Court ruling on the ACA’s Medicaid provision, states were given the choice to opt in or out of expansion, rather than being mandated to participate (4). As a result, states were divided between early adopters of Medicaid expansion and those that opted not to expand. This division created a natural experiment, allowing researchers to study the effects of Medicaid expansion on healthcare access and affordability (5).

Prior evaluations consistently show that Medicaid expansion was linked to improved insurance uptake, especially within economically disadvantaged and minoritized populations (6,7). Furthermore, evidence suggests that expansion has reduced disparities in access to care and financial barriers, such as delaying or forgoing medical care due to cost (8). More recent evaluations have reinforced these findings. For instance, a 2025 study estimated that Medicaid expansion led to more than 27,000 lives saved between 2010 and 2022 (9), while another study reported a mortality reduction of over 30 deaths per 100,000 person-years in expansion states (10). Additionally, new research has documented income increases and financial relief among low-income adults following expansion (11). However, these studies have largely emphasized mortality, income, and administrative outcomes, with limited attention to patient-reported affordability barriers.

Despite the substantial body of research, most evaluations focus on clinical outcomes or disease-specific screenings, such as cancer or diabetes screenings (12). Our study aims to extend the literature by focusing on two broader proxy measures of healthcare access: (1) health insurance coverage, and (2) cost-related barriers to care. By doing so, we address whether Medicaid expansion not only increased coverage but also translated into improved access to necessary healthcare services.

Using the Behavioral Risk Factor Surveillance System (BRFSS) data from 2012, 2013, 2015, 2016, and 2018, this study employs a Difference-in-Differences (DiD) approach to isolate the impact of Medicaid expansion.

## Research Questions and Hypotheses

### Research Question 1

Did Medicaid expansion under the Affordable Care Act increase the likelihood of having health insurance coverage among U.S. adults?

### Hypothesis 1

Medicaid expansion led to increased health insurance coverage among adults

### Research Question 2

Did Medicaid expansion reduce the likelihood of reporting cost as a barrier to accessing healthcare services?

### Hypothesis 2

Medicaid expansion led to a reduction in cost-related barriers to accessing healthcare.

Understanding the effects of Medicaid expansion is essential, especially as policy debates continue regarding further healthcare reforms and coverage expansions. By focusing on self-reported cost barriers, our study contributes an important perspective that complements mortality and insurance studies with a more direct measure of perceived access. Our findings can provide valuable insights for policymakers, healthcare providers, and public health practitioners working to improve access to care across the U.S.

## METHODS

### Study Design

This study employed a quasi-experimental, Difference-in-Differences (DiD) design to evaluate the impact of Medicaid expansion on insurance coverage and cost barriers to healthcare access. We analyzed publicly accessible data from the Behavioral Risk Factor Surveillance System (BRFSS), a yearly, state-level telephone survey administered by the Centers for Disease Control and Prevention (CDC) that gathers information on health behaviors, chronic conditions, and preventive healthcare utilization. Five survey years were selected for analysis: 2012, 2013 (pre-expansion years), and 2015, 2016, and 2018 (post-expansion years). The year 2014 was excluded to allow for a full year post-policy implementation.

### Study Population

The analytic sample included adults aged 18 years and older residing in the United States. We categorized states according to their Medicaid expansion status as of January 1, 2014, using Kaiser Family Foundation criteria(13). States that adopted the expansion by this date were considered expansion states and these included Arkansas, Arizona, California, Colorado, Connecticut, Delaware, Hawaii, Illinois, Iowa, Kentucky, Maryland, Massachusetts, Michigan, Minnesota, Nevada, New Jersey, New Mexico, New York, North Dakota, Ohio, Oregon, Rhode Island, Vermont, Washington, and West Virginia. Non-expansion states were defined as those that had not implemented Medicaid expansion by early 2014. Respondents missing key variables were excluded from relevant analyses.

### Outcome Variables

Two primary outcomes were analyzed. First, insurance coverage was measured using the BRFSS question on current health insurance status. Responses were recoded into a binary variable: insured (1) and uninsured (0). Second, cost-related barriers to care were assessed based on responses to whether there was a time in the past 12 months when the respondent needed to see a doctor but could not because of cost. Responses were similarly recoded into a binary variable: reported cost barrier (1) versus no cost barrier (0).

### Exposure Variables

The key exposure variables were Medicaid expansion status (expand_state: 1 = expansion, 0 = non-expansion) and time period relative to expansion (post_expansion: 1 = 2014 and later, 0 = 2013 and earlier). The interaction term between expansion status and time period was the primary variable of interest, representing the DiD estimator.

### Statistical Analysis

Descriptive statistics were generated to summarize respondent characteristics by Medicaid expansion status, including means for continuous variables (e.g., age) and proportions for categorical variables (e.g., sex, insurance coverage). Logistic regression models with a logit link were used to estimate odds ratios (ORs) and 95% confidence intervals (CIs) for the outcomes of interest. Separate models were run for insurance coverage and cost barrier outcomes, using a difference-in-differences framework to isolate the effect of Medicaid expansion.

The DiD effect was captured by the interaction term between expansion status and post-expansion period. Odds ratios greater than 1 indicated increased odds of the outcome associated with Medicaid expansion, while odds ratios less than 1 indicated decreased odds.

All analyses were conducted using R version 4.3.2. Statistical significance was assessed at the 0.05 level.

### Ethical Considerations

This study utilized publicly available, de-identified data from the BRFSS, which is administered by the U.S. CDC. All participants provided informed consent at the time of original data collection. As the analysis was conducted using secondary, anonymized data, no additional ethical approval or informed consent was required.

## RESULTS

Table 1 displays the baseline characteristics of BRFSS respondents categorized according to whether their state had expanded Medicaid. Compared to individuals in non-expansion states (n = 1,161,921), those in expansion states (n = 1,107,734) were slightly younger on average (54.5 vs. 54.9 years; *p* < 0.001). A higher proportion of respondents in expansion states were female (56.6% vs. 56.0%; *p* < 0.001). Notably, insurance coverage was higher in expansion states (92.2%) than in non-expansion states (89.5%; *p* < 0.001), while the percentage reporting cost-related barriers to care was significantly lower (8.9% vs. 10.4%; *p* < 0.001).

**Table 1.** Demographic and Health Coverage Characteristics by Medicaid Expansion Status.

| **Characteristic** | **Non-Expansion States**<br>**(n=1,161,921)** | **Expansion States**<br>**(n=1,107,734)** | **p-**<br>**value** |
| --- | --- | --- | --- |
| Age (mean ± SD) | 54.9 ± 16.5 | 54.5 ± 18.1 | <0.001 |
| Sex (female) | 56.0% | 56.6% | <0.001 |
| Insurance coverage (insured) | 89.5% | 92.2% | <0.001 |
| Cost barrier to care (reporting cost<br>barrier) | 10.4% | 8.9% | <0.001 |

Figure 1 illustrates trends in adult health insurance coverage from 2012 to 2018, comparing states based on Medicaid expansion status. After the 2014 expansion, insurance rates rose significantly in expansion states, peaking around 2016, while non-expansion states showed smaller or stagnant gains.

**Figure 1.**
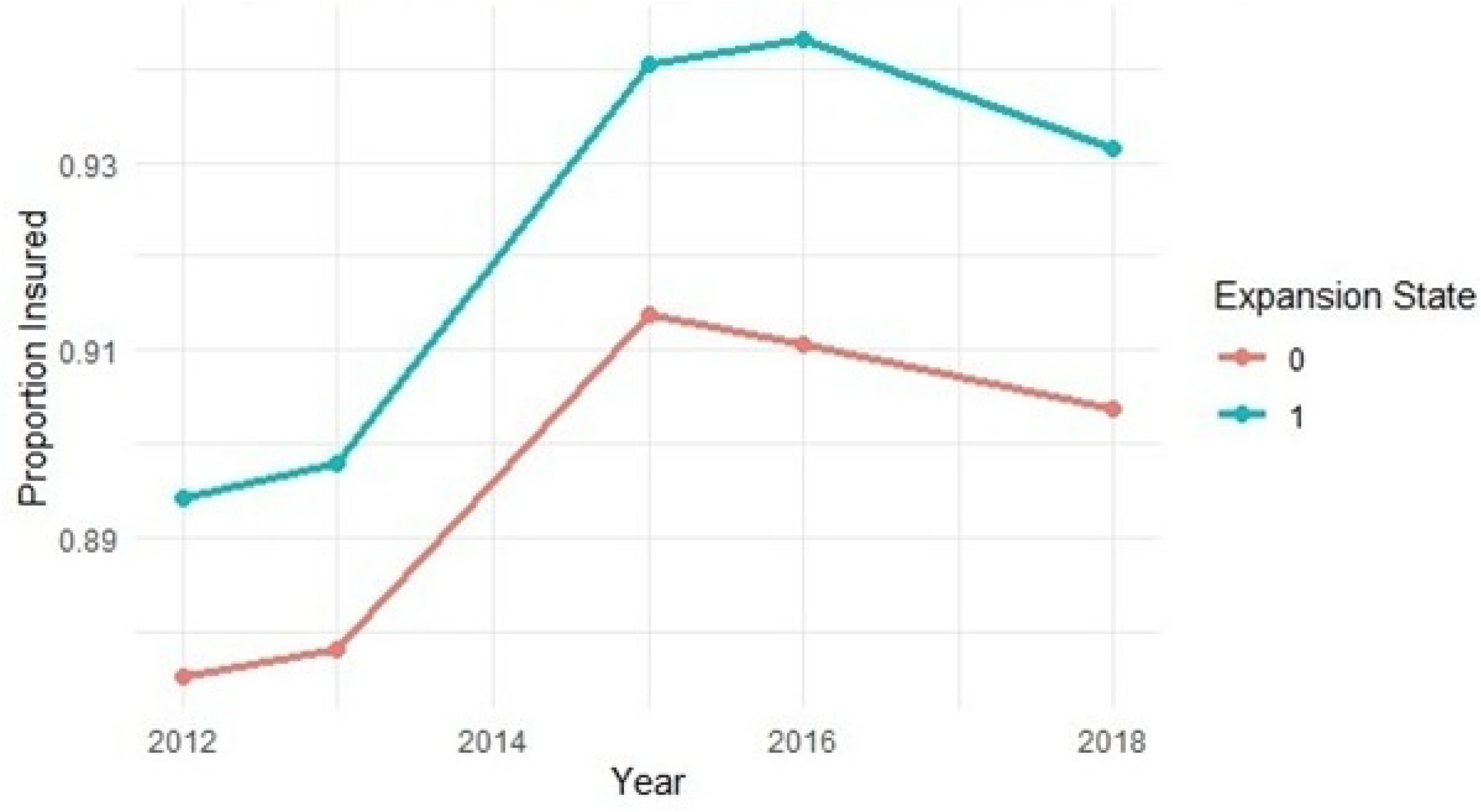
Trend in Health Insurance Coverage by Medicaid Expansion Status, 2012–2018

Figure 2 depicts trends in the percentage of adults who reported cost as a barrier to accessing healthcare over time. Expansion states experienced a sharp decline in cost barriers following Medicaid expansion in 2014, with sustained improvements through 2018. Meanwhile, non-expansion states showed only slight reductions, underscoring the role of Medicaid expansion in reducing financial obstacles to healthcare.

**Figure 2.**
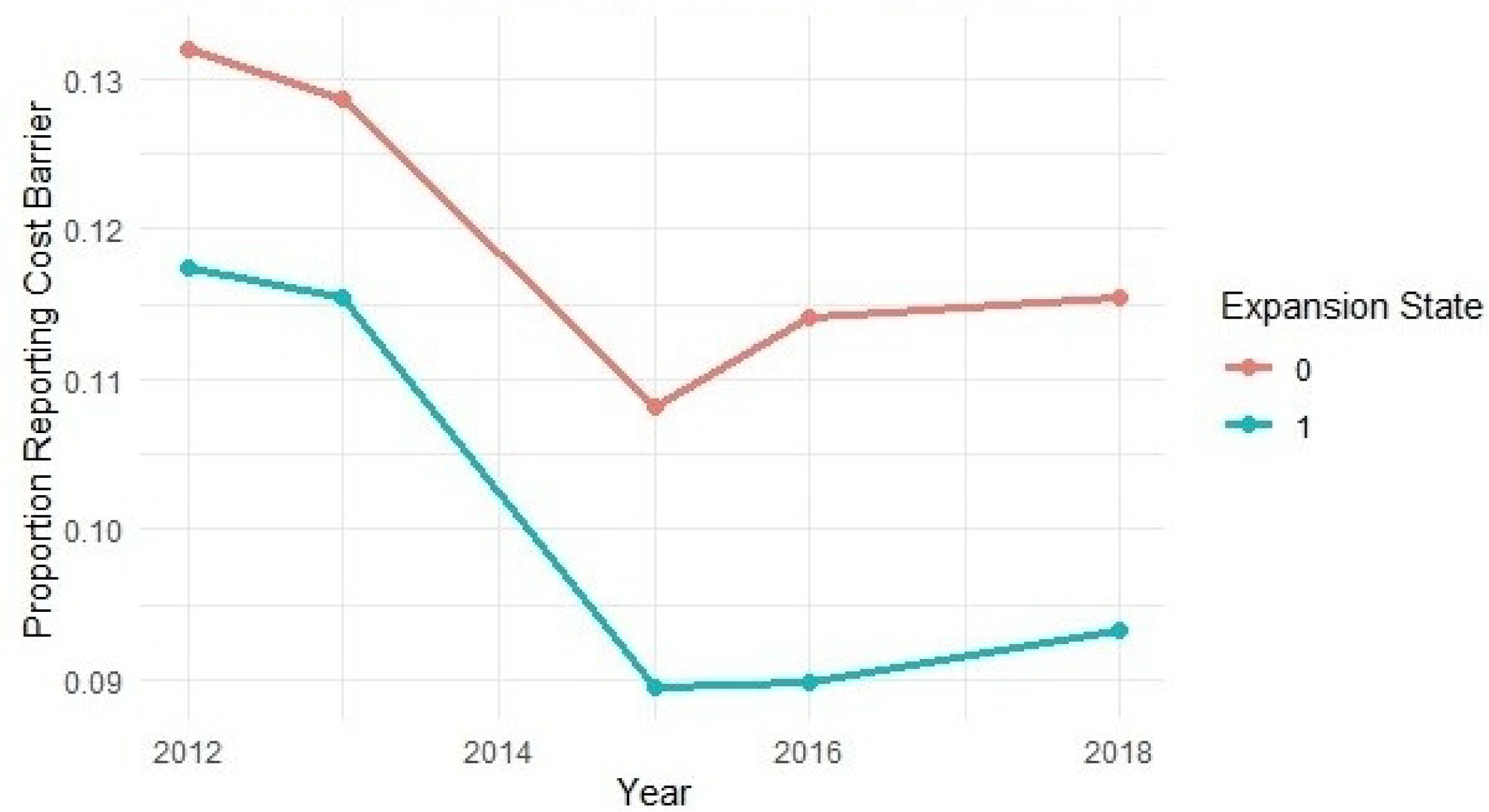
Trend in Cost-Related Barriers to Care by Medicaid Expansion Status, 2012– 2018

**Figure 3.**
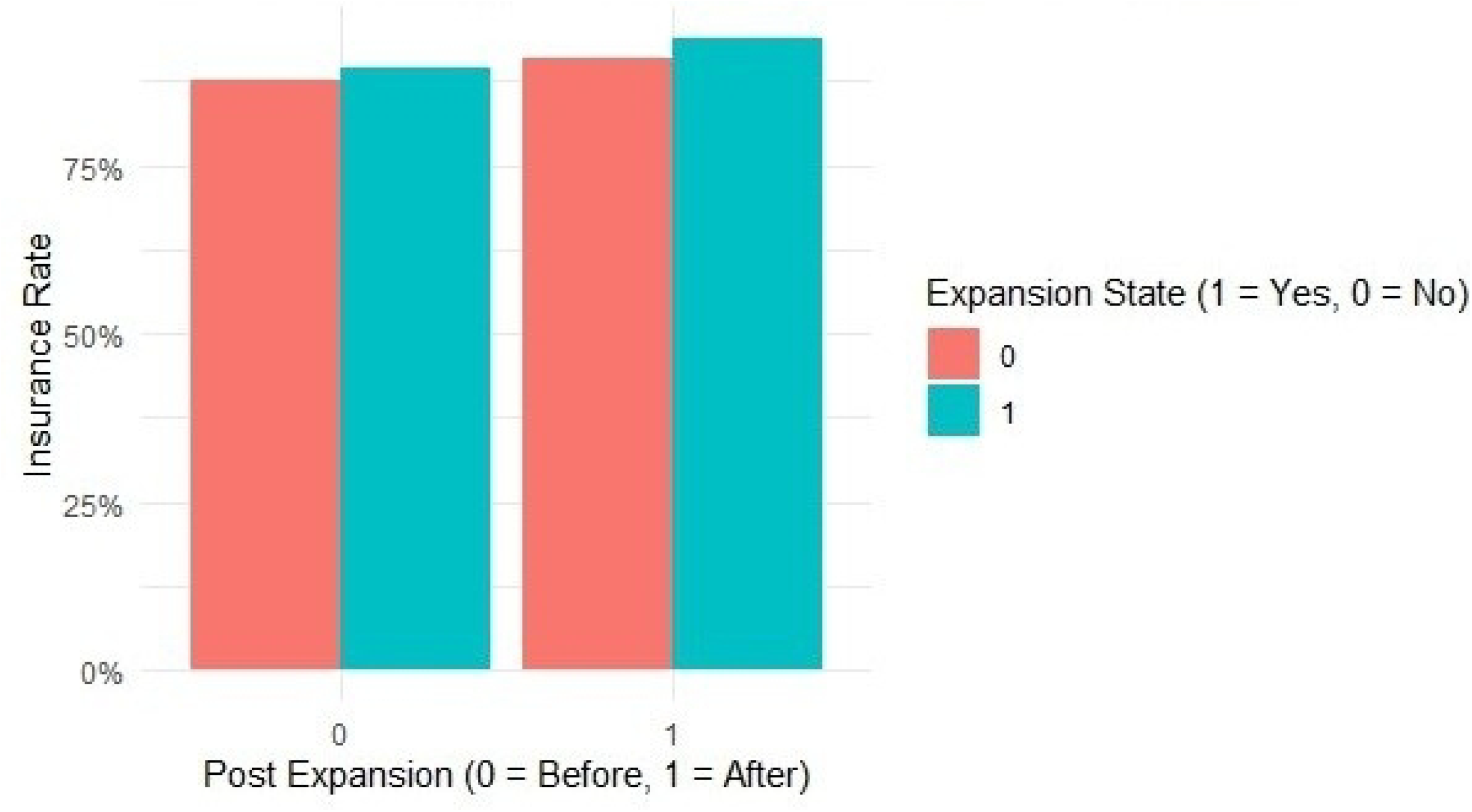
Cost-Related Barriers to Care Before and After Medicaid Expansion

**Figure 4.**
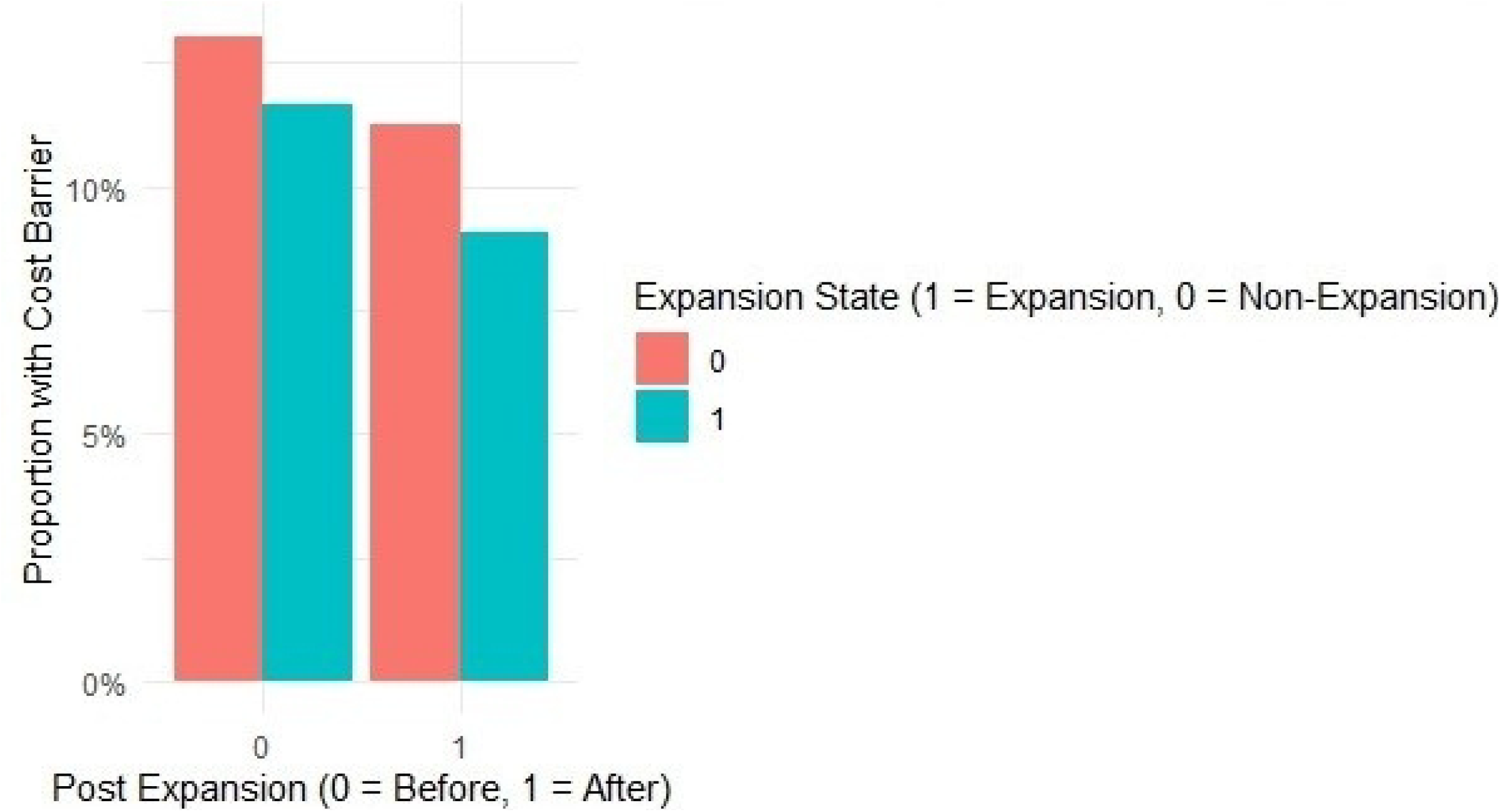
Cost-Related Barriers to Care Before and After Medicaid Expansion

Table 2 reports findings from logistic regression models evaluating how Medicaid expansion affected self-reported financial obstacles to care access. Individuals in expansion states had 21% higher odds of being insured overall, and the post-expansion period was associated with a 41% increase in odds of insurance. The significant interaction term (OR = 1.25; 95% CI: 1.22–1.28) indicates that the combined effect of being in an expansion state after 2014 led to a 25% increase in insurance odds relative to non-expansion states.

**Table 2.** Association Between Medicaid Expansion and Insurance.

| **Predictor** | **OR (95% CI)** | **p-value** |
| --- | --- | --- |
| expand_state | 1.21 (1.19, 1.23) | <0.001 |
| post_expansion | 1.41 (1.40, 1.43) | <0.001 |
| expand_state × post_expansion | 1.25 (1.22, 1.28) | <0.001 |

Table 3 shows the results of a logistic regression assessing the association between Medicaid expansion and reported cost-related barriers to accessing care. Living in an expansion state was associated with 12% lower odds of reporting cost-related barriers (OR = 0.88; 95% CI: 0.87– 0.89), and the post-expansion period independently reduced the odds by 15% (OR = 0.85; 95% CI: 0.84–0.86). The interaction term (OR = 0.90; 95% CI: 0.89–0.91) indicates an additional 10% reduction in cost barriers in expansion states after Medicaid expansion implementation.

**Table 3.** Association Between Medicaid Expansion and Cost-Related Barriers to Care.

| Predictor | OR (95% CI) | p-value |
| --- | --- | --- |
| expand_state | 0.88 (0.87, 0.89) | <0.001 |
| post_expansion | 0.85 (0.84, 0.86) | <0.001 |
| expand_state × post_expansion | 0.90 (0.89, 0.91) | <0.001 |

**Table 4.** Year-Specific Odds Ratios for Insurance Coverage in Expansion vs. Non-Expansion States.

| Year | OR | 95% CI | p-value |
| --- | --- | --- | --- |
| 2012 | 1.21 | 1.18 - 1.23 | <0.001 |
| 2013 | 1.22 | 1.20 - 1.25 | <0.001 |
| 2015 | 1.49 | 1.46 - 1.53 | <0.001 |
| 2016 | 1.63 | 1.60 - 1.67 | <0.001 |
| 2018 | 1.45 | 1.42 - 1.48 | <0.001 |

Annual odds ratios are presented to compare insurance coverage among adults residing in Medicaid expansion versus non-expansion states over the study period. Across all the years analyzed, individuals in expansion states consistently had significantly higher odds of insurance coverage. The difference widened after the 2014 expansion, peaking in 2016 (OR = 1.63; 95% CI: 1.60–1.67), indicating sustained gains in coverage post-implementation of the ACA Medicaid provision.

## DISCUSSION

This study demonstrates that Medicaid expansion under the Affordable Care Act (ACA) significantly increased insurance coverage and reduced cost-related barriers to accessing healthcare. The use of a Difference-in-Differences (DiD) design allowed for a robust estimation of the policy’s effect by comparing trends in expansion versus non-expansion states before and after the 2014 implementation of Medicaid expansion. By isolating the interaction term, this approach captures the causal impact of the policy beyond general time trends or ACA-wide effects.

The results align with previous research that has consistently shown increases in insurance coverage following Medicaid expansion (14) Another study found that expansion states experienced larger gains in coverage compared to non-expansion states, particularly among low-income adults (5). Parallel findings have documented significant declines in the uninsured population, further supporting the policy’s positive impact. Our findings extend this evidence by showing not only higher odds of insurance coverage in expansion states post-expansion (OR: 1.25; 95% CI: 1.23–1.28) but also year-over-year increases in odds ratios from 2012 through 2016, peaking shortly after implementation.

In addition to insurance coverage, the study adds to a growing body of literature evaluating access barriers, particularly cost-related delays in care. We observed that Medicaid expansion was associated with reduced odds of reporting cost as a barrier to care (OR: 0.90; 95% CI: 0.89– 0.91), suggesting that expansion efforts alleviated financial obstacles for many. This is consistent with findings from the Oregon Health Insurance Experiment, which demonstrated reduced out-of-pocket expenditures and financial strain among Medicaid recipients (15). Similarly, a 2025 study found that Medicaid expansion was associated with an 11.3 percentage point reduction in cost-related access barriers and a 13.4 percentage point increase in insurance coverage among low-income parents, reinforcing the broader population-level benefits of expansion (16). Importantly, other studies have found that these financial improvements may also translate to better health outcomes; for instance, Miller and Wherry reported a reduction in mortality associated with Medicaid expansion, particularly among low-income adults (17).

Despite these improvements, it is important to note that some cost barriers persisted even in expansion states, pointing to the multifactorial nature of access issues. Insurance alone may not eliminate all financial hurdles, especially when considering premiums, co-pays, or gaps in provider availability. Moreover, states varied in their implementation timelines, eligibility thresholds, and outreach efforts, which could contribute to heterogeneity in outcomes (18). These mixed results echo findings from a systematic review by Mazurenko et al (19), which found that while Medicaid expansion improved access and health outcomes overall, challenges remain in ensuring equitable care and consistent implementation across states.

### Strengths And Limitations

This study has several notable strengths. First, it utilizes data from the Behavioral Risk Factor Surveillance System (BRFSS), a large and nationally representative survey of U.S. adults, which enhances the generalizability of findings. Second, the use of a difference-in-differences (DiD) approach leverages a natural quasi-experimental setting created by the staggered adoption of Medicaid expansion across states, allowing for more robust causal inference.

However, the study also has important limitations. The reliance on self-reported survey data introduces the potential for recall and social desirability bias, particularly regarding sensitive measures such as insurance status and cost-related barriers to care. Additionally, although the DiD method helps to control for time-invariant unobserved confounders, residual confounding from unmeasured time-varying factors may still bias results. Finally, states differed not only in whether they adopted Medicaid expansion but also in the timing and specific implementation strategies, which may limit the comparability of expansion effects across states.

## CONCLUSION

The findings from this study suggest that expanding Medicaid under the Affordable Care Act was associated with notable improvements in health insurance coverage and a reduction in financial barriers to care. States that implemented the expansion observed larger increases in coverage rates and more pronounced decreases in cost-related challenges than those that did not. These results reinforce the value of Medicaid expansion as a policy tool to promote health equity and improve access among underserved populations.

## Data Availability

The datasets analyzed during the current study are available from the CDC BRFSS data repository: https://www.cdc.gov/brfss/annual_data/annual_data.htm

## Declarations

### Ethics approval and consent to participate

This study is a secondary analysis of publicly available, de-identified data from the Behavioral Risk Factor Surveillance System (BRFSS), collected by the Centers for Disease Control and Prevention (CDC). The dataset is anonymized and publicly accessible and therefore exempt from institutional review board (IRB) approval. No additional ethical approval was required.

### Consent for publication

Not applicable.

### Authors’ contributions

NN conceptualized the study, conducted the data analysis, and wrote the original draft. BC, HM and LS contributed to review and editing of the manuscript. All authors read and approved of the final manuscript.

## Acknowledgements

The authors acknowledge the Centers for Disease Control and Prevention (CDC) for providing access to the BRFSS data used in this analysis. The authors would also like to thank Maisy Knight, Georgetown University Graduate School of arts and Sciences, for her valuable support in reviewing and providing feedback on earlier versions of this manuscript.

